# Monitoring life expectancy levels during the COVID-19 pandemic: Example of the unequal impact in Spanish regions

**DOI:** 10.1101/2020.06.03.20120972

**Authors:** Sergi Trias-Llimós, Tim Riffe, Usama Bilal

## Abstract

**Background:** To provide an interpretable summary of the impact on mortality of the COVID-19 pandemic we estimate weekly and annual life expectancies at birth in Spain and its regions.

**Methods:** We used daily death count data from the Spanish MoMo, and death counts from 2018, and population on 1 July, 2019 by region (CCAA), age groups, and sex from the Spanish National Statistics Institute. We estimated weekly and annual (2019 and 2020*, the shifted annual calendar period up to June 14th 2020) life expectancies at birth as well as their differences with respect to 2019.

**Results:** Weekly life expectancies at birth in Spain were lower in weeks 11-20, 2020 compared to the same weeks in 2019. This drop in weekly life expectancy was especially strong in weeks 13 and 14 (March 23^rd^ to April 5^th^), with national declines ranging between 6.1 and 7.6 years and maximum regional weekly declines of up to 15 years in Madrid. Annual life expectancy differences between 2019 and 2020 also reflected an overall drop in annual life expectancy of 0.8 years for both men and women. These drops ranged between 0 years in several regions (e.g. Canary and Balearic Islands) to 2.7 years among men in Madrid.

**Conclusions:** Life expectancy is an easy to interpret measure for understanding the heterogeneity of mortality patterns across Spanish regions. Weekly and annual life expectancy are sensitive useful indicators for understanding disparities and communicating the gravity of the situation because differences are expressed in intuitive year units.

**Key messages:**

- Weekly and annual updated life expectancy are valuable indicators of the health impacts of the pandemic in populations.
- The impact of the COVID-19 pandemic in Spain has been severe and highly heterogeneous, with weekly life expectancy falls of up to 15 years in Madrid, and with annual life expectancy falls ranging between 0 and 2.7 years.
- Our results for Spain provide important insights into the magnitude of the pandemic in mortality levels across regions and are easy to interpret and compare.

## Introduction

The COVID-19 pandemic is causing substantial increases in mortality in several populations worldwide. According to WHO, by June 22^nd^, 2020 over 465,000 confirmed COVID-19 deaths occurred worldwide (1). Spain has been one of the most affected countries with more than 28,000 deaths with laboratory confirmation of COVID-19 (1). Nonetheless, both the official number of COVID-19 cases and deaths, and the results from a recent seroprevalence study reveal important differences across Spanish regions (2,3).

Beyond the official death toll statistics, the COVID-19 pandemic has been associated with net increases in mortality in several populations (4), which could owe to a combination of factors. The COVID-19 pandemic used an unprecedented amount of health service resources, including intensive care unit beds, and strong preventive health measures in hospitals. This has put health systems in struggling and challenging situations (5), and thus potentially leading to increases in morbidity and mortality indirectly related to COVID-19. For example, some excess mortality could result from healthcare avoidance, from a delay in treatment, or from insufficient care for other urgent conditions resulting from a reduced capacity to treat other medical emergencies. Other kinds of mortality may be temporarily reduced, such as deaths from acute respiratory conditions related to air pollution, or traffic accidents, but those mortality reductions may be outweighed by excesses. While total excess mortality and total number of COVID-19 deaths provide some measure of the impact of the pandemic within populations, their interpretation is not always straightforward, and comparisons between populations can be challenging.

Few studies assessing the impact of the pandemic on mortality have reported life expectancy estimates. Life expectancy is a summary index of mortality that is easy to interpret, but that is also subject to common misunderstandings. The most literal understanding of life expectancy is as the expected average length of life that would follow in the long run if a given set of mortality conditions were held fixed. The most common misunderstanding is to treat it as a forecasted expectancy. In monitoring mortality patterns in non-crisis times, annual life expectancy is a standard indicator because it accounts for differences in age-specific mortality, and it is expressed in intuitive year units, making it easy to grasp and compare across populations and over time.

The current pandemic, with fast mortality increases in specific weeks, makes it useful to shorten the calendar reference period of mortality rates in order to measure life expectancy changes. When calculated over short time intervals in this way, life expectancy should be understood as an annualized summary index of mortality. This index is much more volatile than standard period life expectancy calculated over a year, and it should be reported alongside traditional calendar-year life expectancy measures, possibly with shifting calendar reference windows. A few studies that have documented life expectancy declines have found notable declines in annual life expectancy in highly affected Spanish and Italian regions, Madrid and Bergamo (6,7), and declines in weekly life expectancy in Sweden (8).

The objective of this study is to estimate the impact of the COVID-19 pandemic by estimating both weekly and annual life expectancies in Spain and its 17 regions.

## Methods

### Settings

We estimated life expectancy at birth by sex in Spain and its 17 regions (*comunidades autónomas*) in two time frames: i) weekly life expectancy at birth from week 1 in 2019 until the most recently available weekly data (week 24, June 8-14); and ii) annual life expectancy for both 2019 and the shifted annual reference period up to June14^th^, 2020 (referred as 2020*). Due to small population sizes, we excluded the Spanish cities of Ceuta and Melilla on mainland Africa from the analyses.

### Data

Data sources included daily age- (<65, 65-75, 75+) and sex-specific death counts data from the Spanish *Sistema de vigilancia de la mortalidad diaria* (Daily Mortality Monitoring System, MoMo, updated May 28^th^) covering ∼93% of the population (9). We also used sex- and age-specific (5-year age groups) death counts by region in 2018 from Spanish National Statistics Institute (INE) (10), and population estimates from July 1^st^, 2019 from INE (11). To compare changes in life expectancy with the number of infected people, we obtained data on the region-level seroprevalence of IgG against SARS-COV-2 from the seroprevalence study in Spain carried out in over 60,000 individuals between April 27 and May 11 2020 (2).

### Methods

We conducted our analysis in five steps. First, we grouped daily death counts into weeks. Second, we redistributed the MoMo death counts from broad age groups (<65, 65-75, and 75+) into 5-year age groups using 2018 death counts from the INE as a proportional standard. Third, we estimated age-specific death rates for each population group using the 2019 mid-year population as the denominator for annual estimates, and the population divided by (365/7) as denominator for weekly estimates. Fourth, both weekly and annual life expectancies were estimated using conventional life table techniques, and 95% confidence intervals were estimated based on the 2.5 and 97.5th percentiles of 1,000 random binomial deviates (12,13). Finally, we derived the expected variation in annual life expectancy between 2019 and 2020* by subtracting life expectancy at birth in 2019 from life expectancy at birth in 2020*.

To assess the robustness of our estimates we visually inspected the associations between the annual life expectancy variation and the IgG anti SARS-Cov2 prevalence by region and sex.

## Results

In Spain, weekly life expectancies at birth for weeks 11-20 2020 were lower than those from the same weeks in 2019 (**Figure 1**, and Appendix I **Figure S1**). This drop in weekly life expectancy was especially strong in weeks 13 and 14 (23 March to 5 April), with national declines ranging between 6.1 and 7.6 years. This drop in weekly life expectancy was heterogeneous across Spanish regions. Madrid experienced the most important drop, which ranged between 11.2 and 14.8 years in weeks 13-14 for both men and women. Catalonia also experienced important drops, ranging between 8.4 and 9.4 years, along with Castile-La Mancha and Castile and Leon (Appendix I **Figure S2**). North-western regions of Galicia and Asturias, along with Murcia and the Canary Islands, did not experience major disruptions in weekly life expectancy. From weeks 21 (May 18-24) onwards weekly life expectancy in 2020 has been at levels close to 2019 in Spain and in most of its regions, including Madrid and Catalonia.

**Figure 1.**
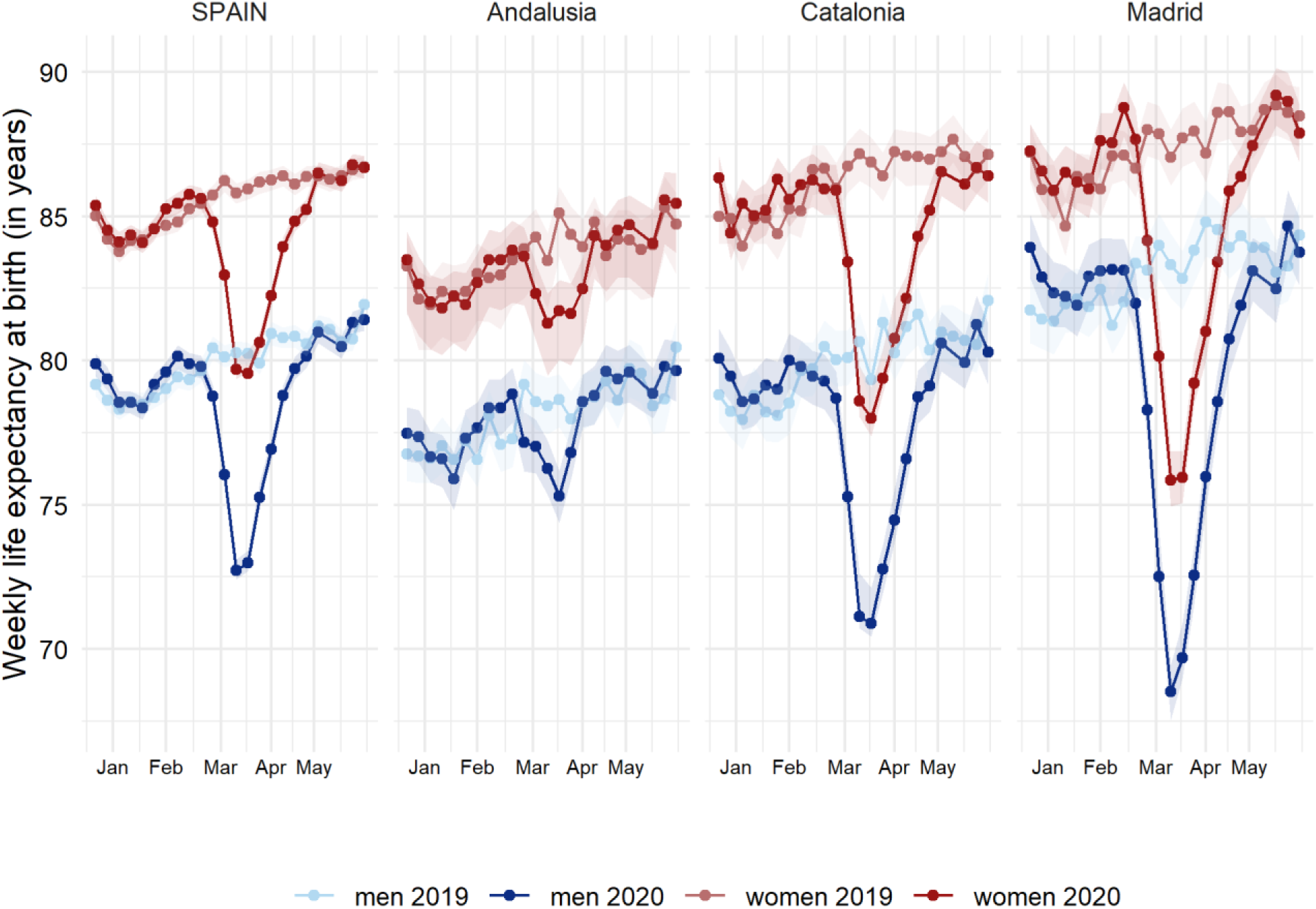
Weekly life expectancy at birth (with 95% confidence intervals) in Spain and three selected regions (Andalusia, Catalonia and Madrid)* by sex (weeks 1-24, 2019 and 2020) * Weekly life expectancies at birth for Spain and its 17 regions can be found in **Figure S1**.

Annual life expectancy differences between 2019 and 2020* also reflected an overall drop in annual life expectancy of 0.8 years for both men and women (**Figure 2**, and Appendix I **Figure S3**). However, this difference was heterogeneous across regions. Declines were steepest for Madrid, with a loss of 2.7 (95%CI: 2.6-2.9) years for men and 2.0 (1.9-2.1) years for women. The least affected regions (Canary and Balearic Islands, Andalusia, and Galicia) show no change in annual life expectancy in 2020* compared to 2019.

**Figure 2.**
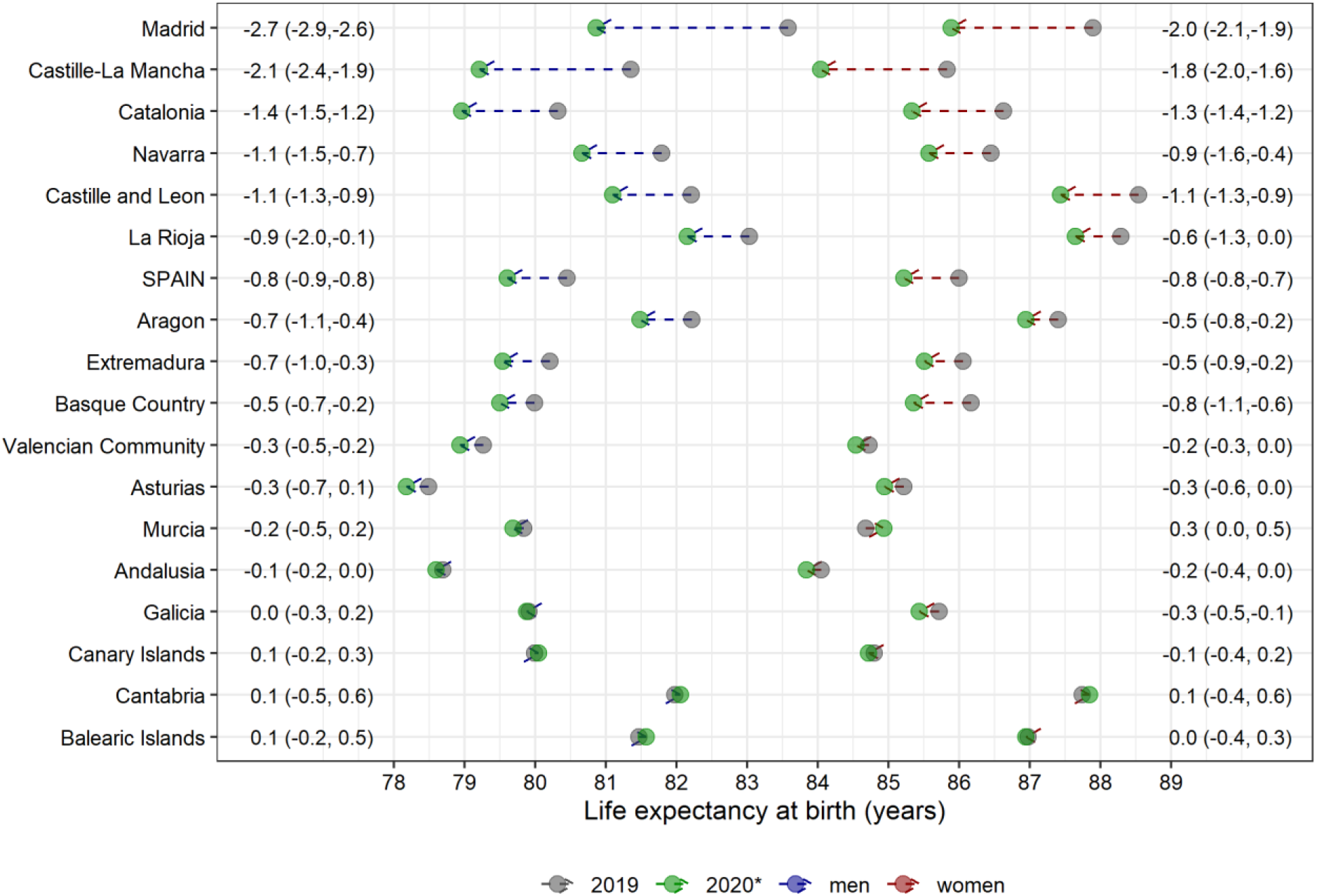
Annual life expectancy at birth in 2019, 2020* and differences between periods for Spain and its 17 regions by sex * Annual life expectancy at birth in 2020* was estimated using death counts from the shifted annual reference period up to 14 June 2020.

Visual inspection on the robustness of our results suggested the observed declines in annual life expectancy to be well aligned with the seroprevalence study in Spain carried out in over 60,000 individuals between April 27 and May 11 2020 (2) (**Figure 3**). The Pearson correlation coefficient was 0.96 and 0.91 for men and women, respectively.

**Figure 3.**
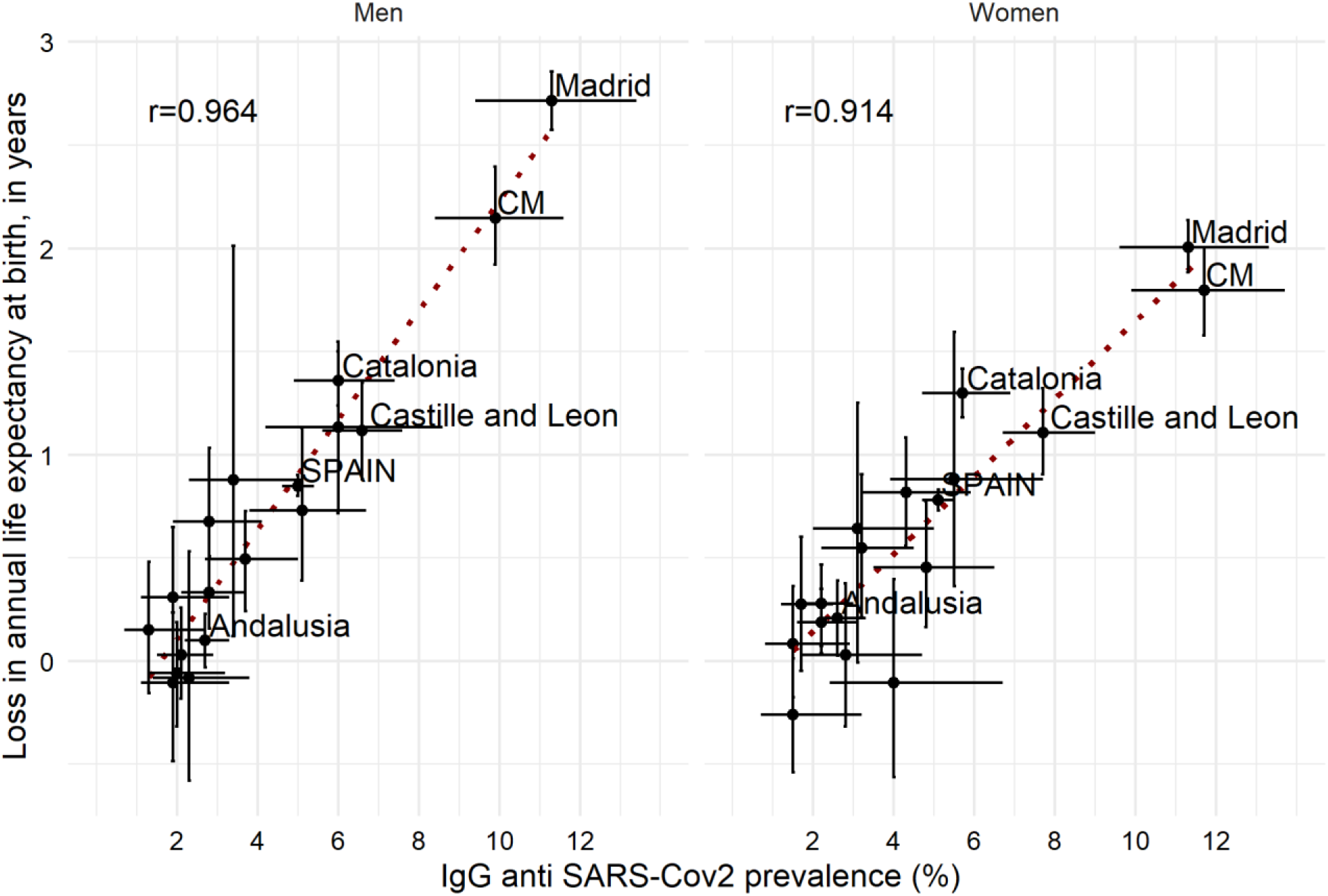
Associations between loss in annual life expectancy at birth between 2019 and 2020* and IgG anti SARS-Cov2 prevalence for Spain and its 17 regions by sex (95% CI). * Annual life expectancy at birth in 2020* was estimated using data from the one year window that closes out June 14, 2020. **CM stands for Castille-La Mancha.

## Discussion

In this study, we have documented for the first time the impact of the COVID-19 pandemic on weekly and annual life expectancies at birth at the regional level in Spain, one of the most severely affected countries in the world. The heterogeneity of the pandemic across Spain is reflected in wide differences between regions, with some of the most affected regions losing up to 10-15 years of weekly life expectancy and more than 2 years in annual life expectancy, whereas other regions’ life expectancy was hardly affected. These changes in life expectancy correlated strongly with the prevalence of antibodies against SARS-COV-2.

Our estimates of the 2-3 years drop in annual life expectancy for Madrid, the most severely hit region, appear slightly lower than similar estimates from the Italian province of Bergamo up to April 30th 2020 (7). However, this comparison should be taken cautiously as the time frame and pandemic timing differed between studies, and life expectancy was compared to 2017 in the Bergamo study whereas we compared with 2019 levels. Our estimates of a 2-3 years of drop in life expectancy in the most affected weeks in some of the least affected regions (e.g. Andalucia or Cantabria) (see Appendix I, **Figure S4**) are comparable with a previous study for Sweden (8), in line with the higher impact of COVID-19 in Spain compared to Sweden (1).

There are plausible reasons to think that mortality in the remainder of 2020 may further increase compared to 2019. The epidemic is not yet over at the time of this writing, although none of the Spanish regions are still experiencing excess mortality. Additionally, we cannot rule out a potential second wave of COVID-19 in Spain, and if it happens it may affect regions differently than the first wave. Furthermore, the effects of delayed care of chronic conditions, cumulative anxiety, alcohol consumption, and other factors during the pandemic may contribute to elevated mortality in the coming months. However, mortality may also decrease in the coming months due to the mortality selection of frail individuals which usually occurs after severe flu episodes. That is, the COVID-19 pandemic has been more fatal among elderly individuals with pre-existing health conditions (14,15) as well as individuals residing in nursing homes (15,16). Therefore, it is plausible that some individuals who would have been expected to die in the remaining part of the year have already died, which could lead to mortality reductions in the second half of 2020. Indeed, this seems to occur in the regions of La Rioja and Navarra, where increases in weekly life expectancy are observed in the last week of May and first weeks of June as compared to 2019 (**Figure S1** and **Figure S4**). We do not yet know to what extent such processes might offset one another, and careful monitoring of weekly life expectancy for the remainder of the year should provide answers on the overall impact of the COVID-19 pandemic.

Spain is one of the most affected countries in the first months of the pandemic both in terms of directly related COVID-19 deaths (1), as well as in terms of total excess mortality (4). We estimated the annual life expectancy drop between 2019 and the one year window that closes out June 14, 2020 to be 0.8 years in Spain. In contrast, the average annual increase in life expectancy in Spain increased on average two months per year from 2009 to 2019. Altogether, this suggests that life expectancy drop between observed and expected annual life expectancy in the recent one year window would be around or below one year.

Other populations seem to be as affected or more affected by COVID-19 than Spain. For instance, the UK is at the time of writing the country with the highest relative excess mortality (17), and important geographical inequalities exist as well, with London leading the negative ranking of relative excess mortality (18). Indeed, metropolitan areas with important public transport networks tend to be more affected than other regions within a country, not only in Spain or the UK, but also in Italy, where Lombardy was by far the most affected region (19). Other highly affected metropolitan areas include New York city, where the excess mortality was three-fold higher during seven weeks (20). Other populations, for example Brazil, are somewhat behind European countries in the pandemic, but rapidly experiencing a dramatic death toll increase. If the age distribution of deaths in other populations skews younger, this may lead to stronger impacts on life expectancy. Monitoring weekly and annual life expectancies in the most affected populations would provide clear and understandable information on the impact of the pandemic on mortality at the population level. This information would serve to easily compare the impact across populations.

Our analyses are based on detailed daily death counts data covering 93% of the population. We recognize that this could result in slightly overestimated life expectancies levels, especially for the four regions with real coverage <80% (Aragon, Cantabria, Castile and Leon, and La Rioja). Therefore, life expectancies per se need to be interpreted cautiously. However, the undercoverage of the data used is unlikely to substantially affect our main outcome, the differences between life expectancies (see **Appendix II** for details and for sensitivity analyses exploring the impacts of undercoverage).

The weekly life expectancy estimates presented in this study summarize the intensity of mortality increases (8). Weekly life expectancy is a sensitive, intuitive, and comparable translation of age-standardized excess mortality. Weekly-estimated life expectancies can be compared with standard annual life expectancies and with similar estimates from this or other mortality shocks, but one must be careful not to overinterpret this index as a forecast. For example, our results for Madrid on the weekly life expectancies in weeks 13 and 14 shows a substantial drop of up to 15 years, while the annual life expectancy from May 2019 to May 2020 shows a 2.7 year drop. This is not a provisional estimate of the 2020 life expectancy impact, which would require a forecast of mortality through the end of the calendar year.

In conclusion, the impact of COVID-19 pandemic has been severe and highly heterogeneous in Spain. Weekly and annual updated life expectancy are valuable indicators of the health impacts of the pandemic in populations, and thus should be continuously monitored. Such monitoring efforts should be sustained by up-to-date information on all-cause mortality disaggregated by age and sex. Detailed and updated mortality data should be released by public health agencies and governments worldwide (21,22), requiring an increase of the coverage of electronic vital event reporting and seek to reduce other sources of reporting lags.

## Contributors

The study was conceived by STL, and designed by STL, TR and UB. STL ran the analyses with input from TR and UB. STL drafted the manuscript and all authors participated in critical revision of the manuscript for important intellectual content. TR and UB supervised the study. All authors approved the final manuscript and were responsible for the decision to submit for publication.

## Data Availability

The data and R codes that were used as input for the tables and figures are available here: https://osf.io/fwrk3/

## Appendix I

**Figure S1.**
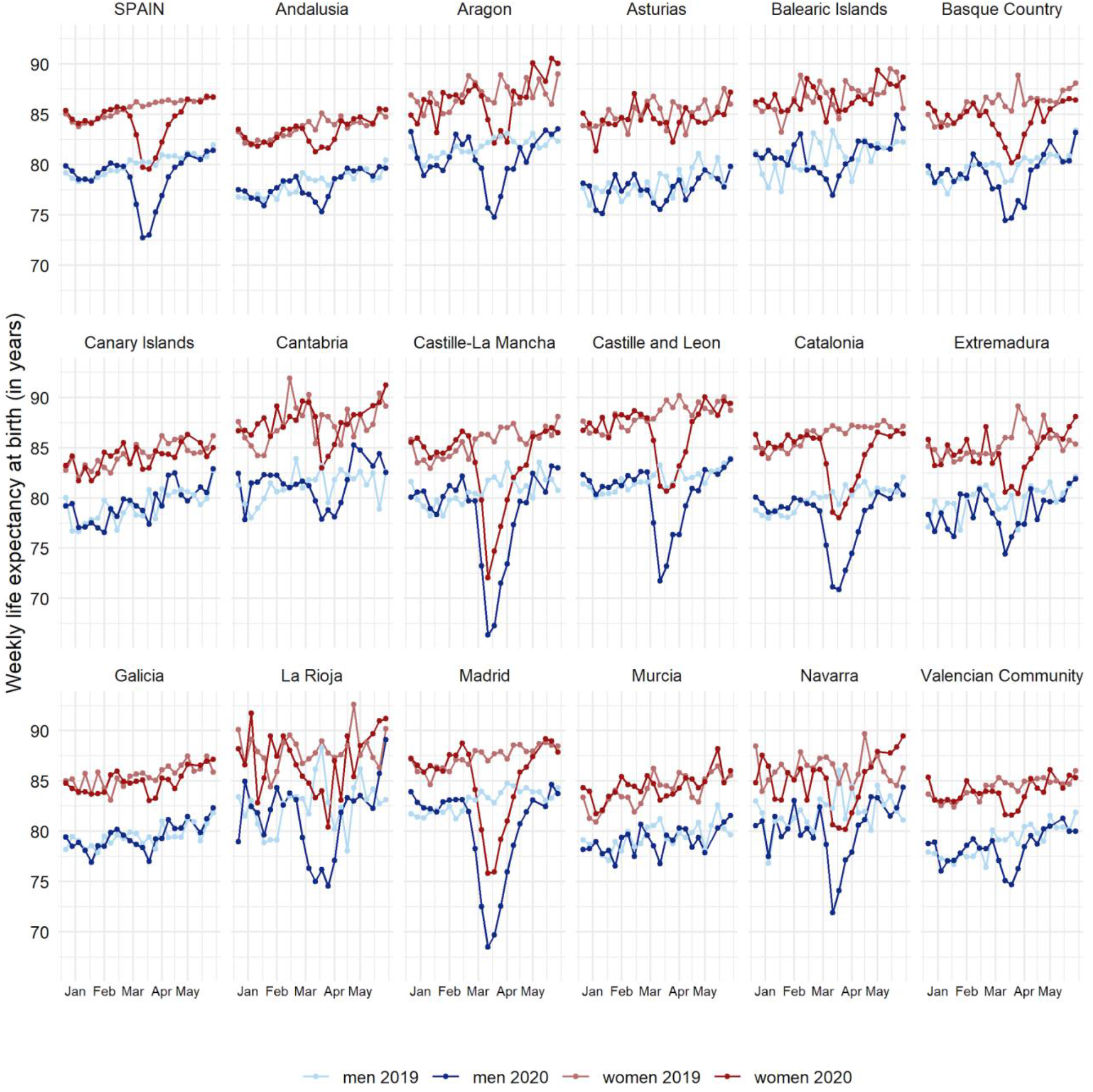
Weekly life expectancy at birth in Spain and its 17 regions by sex (weeks 1-24, 2019 and 2020)

**Figure S2.**
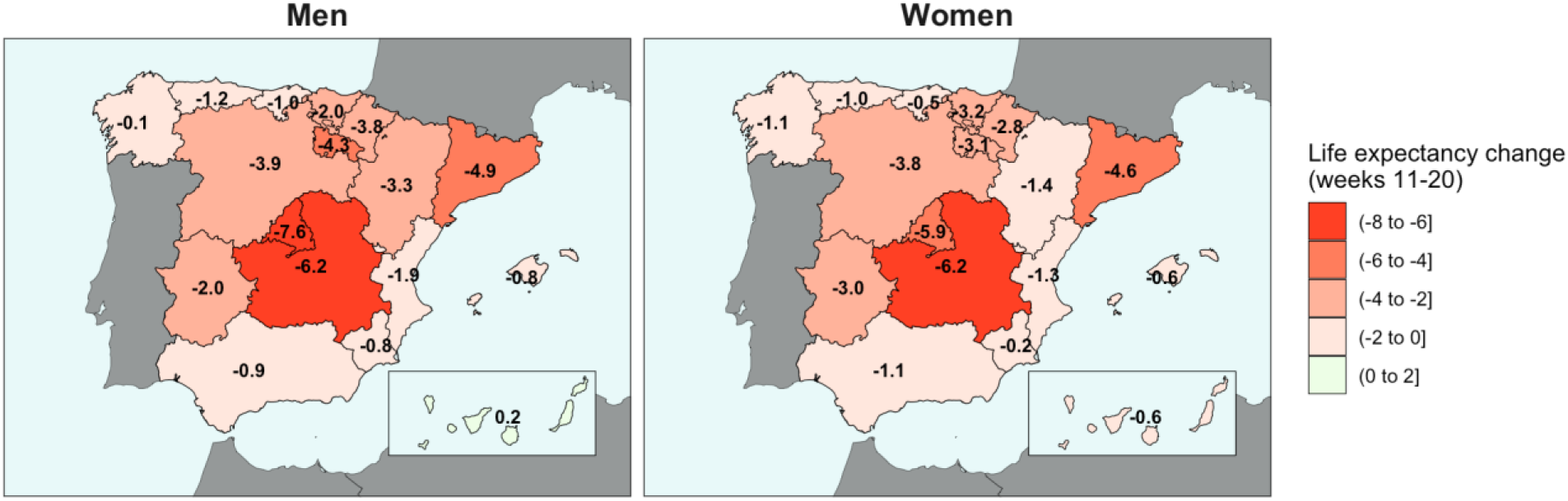
Average weekly life expectancy change in weeks 11-20, 2020 (March 9^th^ till May 17^th^) compared to the corresponding weeks in 2019.

**Figure S3.**
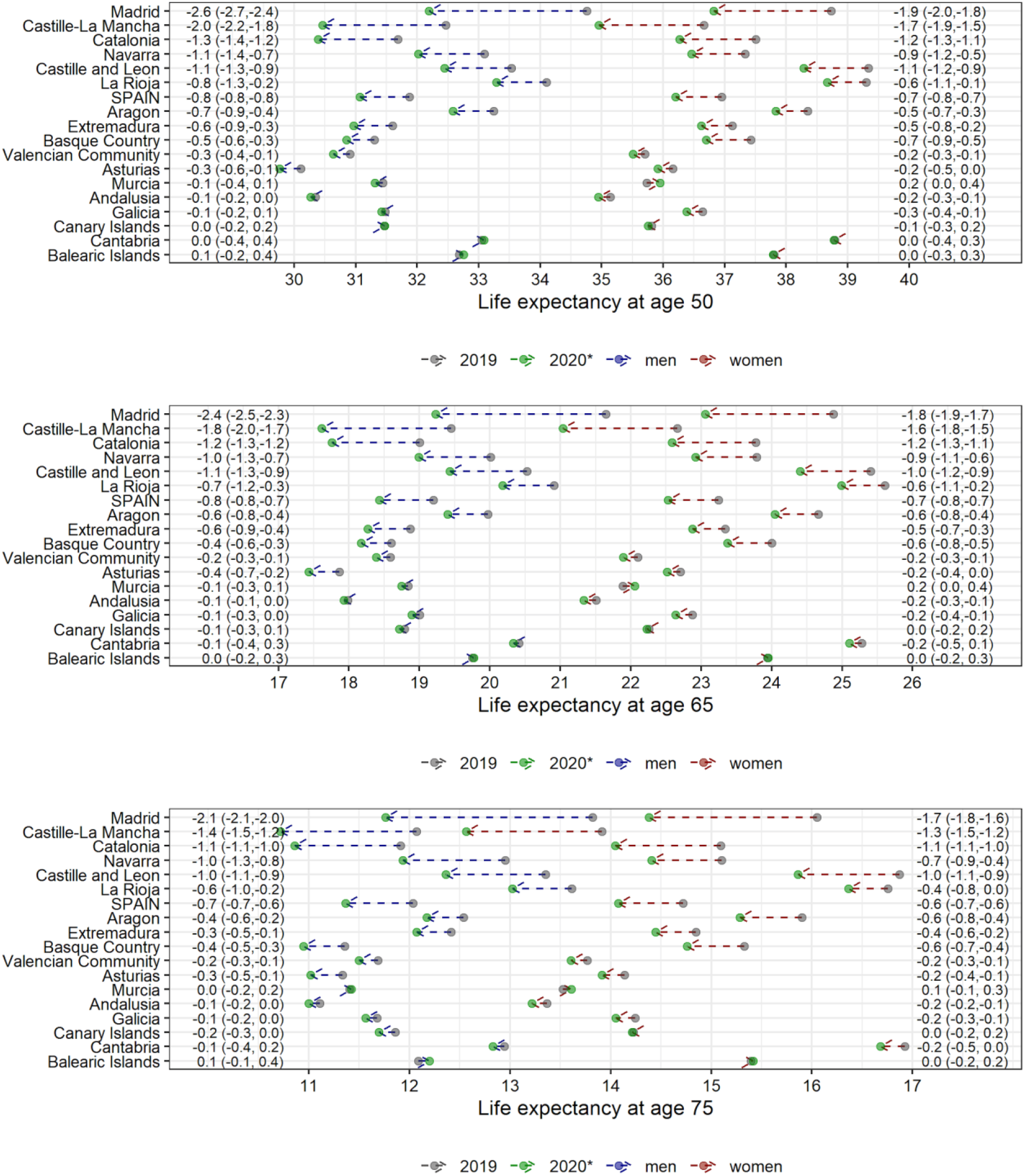
Annual life expectancy at ages 50 and 65 and 75 in 2019, 2020* and differences between periods for Spain and its 17 regions by sex * Annual life expectancy at birth in 2020* was estimated using death counts from the shifted annual reference period up to 14 June 2020.

**Figure S4.**
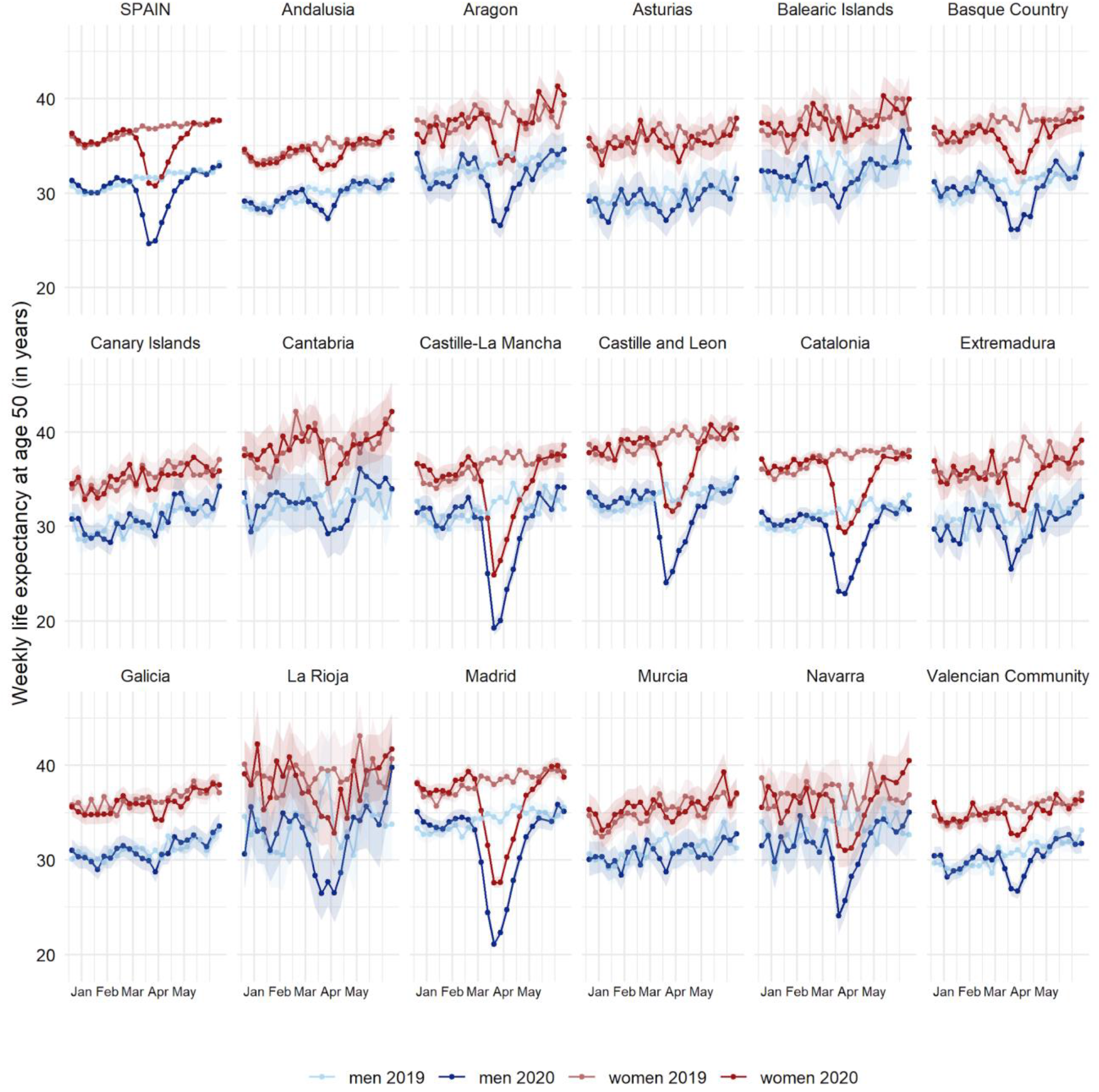
Weekly life expectancy at age 50 (with 95% confidence intervals) in Spain and its 17 regions by sex

## Appendix II: Spanish MoMo coverage and implications for this study

The Spanish daily mortality data that we have used has an overall coverage of ∼93% of the population. We have done several sensitivity analyses to assess the robustness of our results:

1. Comparison between MoMo coverage and estimated coverage based on data from the 1st semester of 2019 suggests that for all regions except Aragon, Cantabria, Castille and Leon, and La Rioja had a real coverage >85%. Ten out of the 17 regions had a coverage >95%. See **Figure 1** for further details.
2. Comparison of estimated annual life expectancies at birth in 2019 (using MoMo data) with those from INE 2018 suggest small differences in estimated life expectancies in 2019 compared to 2018. Regions with real coverage below 85% were the regions where the change in life expectancy is higher. This suggested potential overestimation of life expectancies in the regions with lower coverage. See **Figure 2**.
3. Combining points (1) and (2) we observe a correlation between the real coverage and the differences in life expectancy suggesting regions with low coverage to have potentially higher biases in life expectancy estimates. See **Figure 3**.
4. A sensitivity analysis correcting by undercoverage was done by multiplying death counts by (1/coverage). As this is not advised by MoMo (See here: https://momo.isciii.es/public/momo/dashboard/momo_dashboard.html#documentacion), we used the coverage from the 1st semester of 2019 that we estimated and showed in Appendix II Figure 1. **See Figure 4**.
5. Differences between original annual life expectancy at birth estimates and the corresponding estimates derived from the sensitivity analyses. This figure provides general insights on the extent of life expectancy overestimates. See **Figure 5**.

In conclusion, we aimed at estimating differences in life expectancies during the COVID-19 pandemic. Due to data limitations life expectancy levels may be overestimates, especially for Aragón, Cantabria, Castille and Leon, and la Rioja, and should be interpreted carefully.

However, the undercoverage of the data used is unlikely to substantially affect our main outcome, the differences between life expectancies.

---

**Figure 1.**
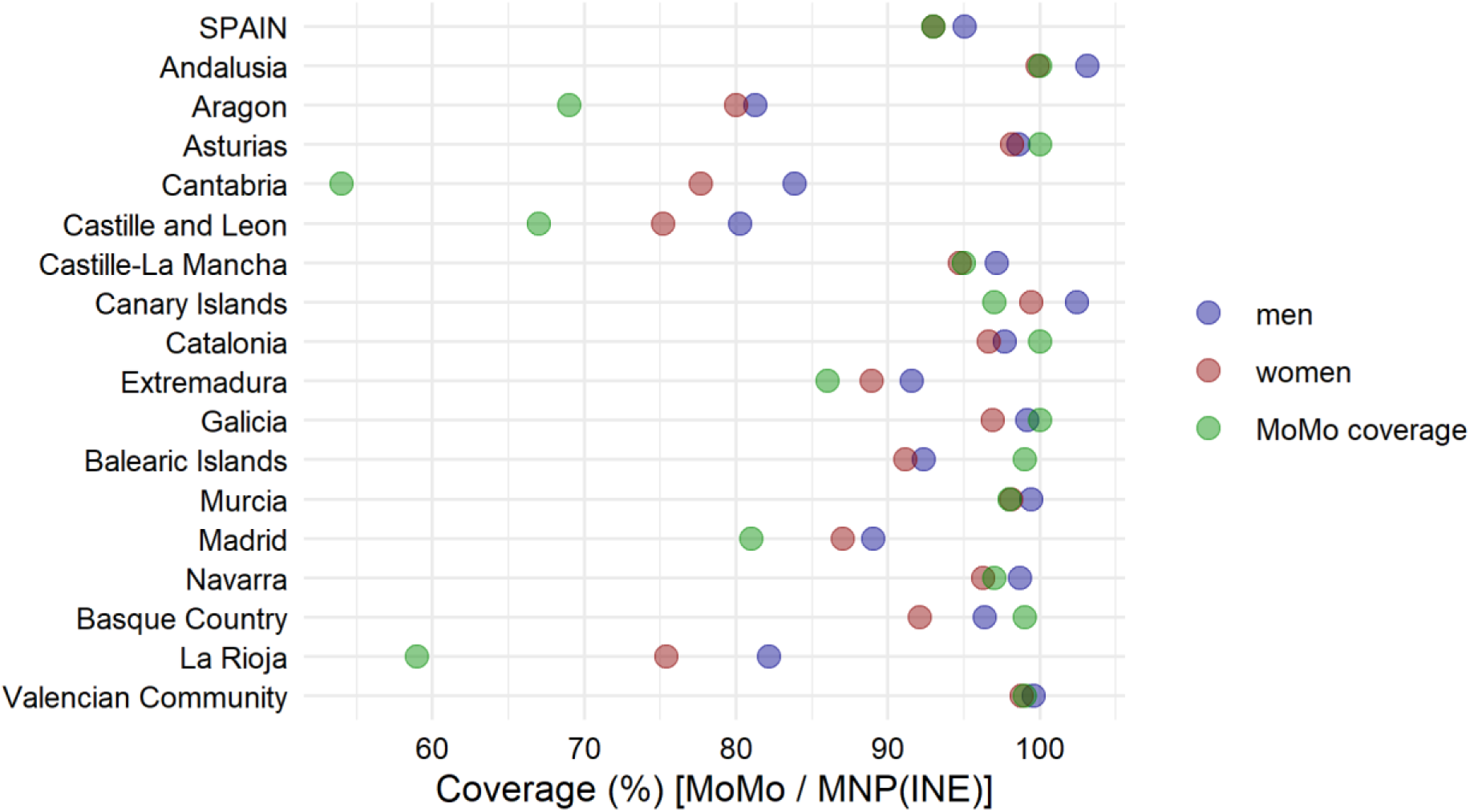
MoMo coverage in semester I 2019 compared with data from “Movimento Natural de la Población” (INE) by region and sex and reported MoMo coverage by region.

**Figure 2.**
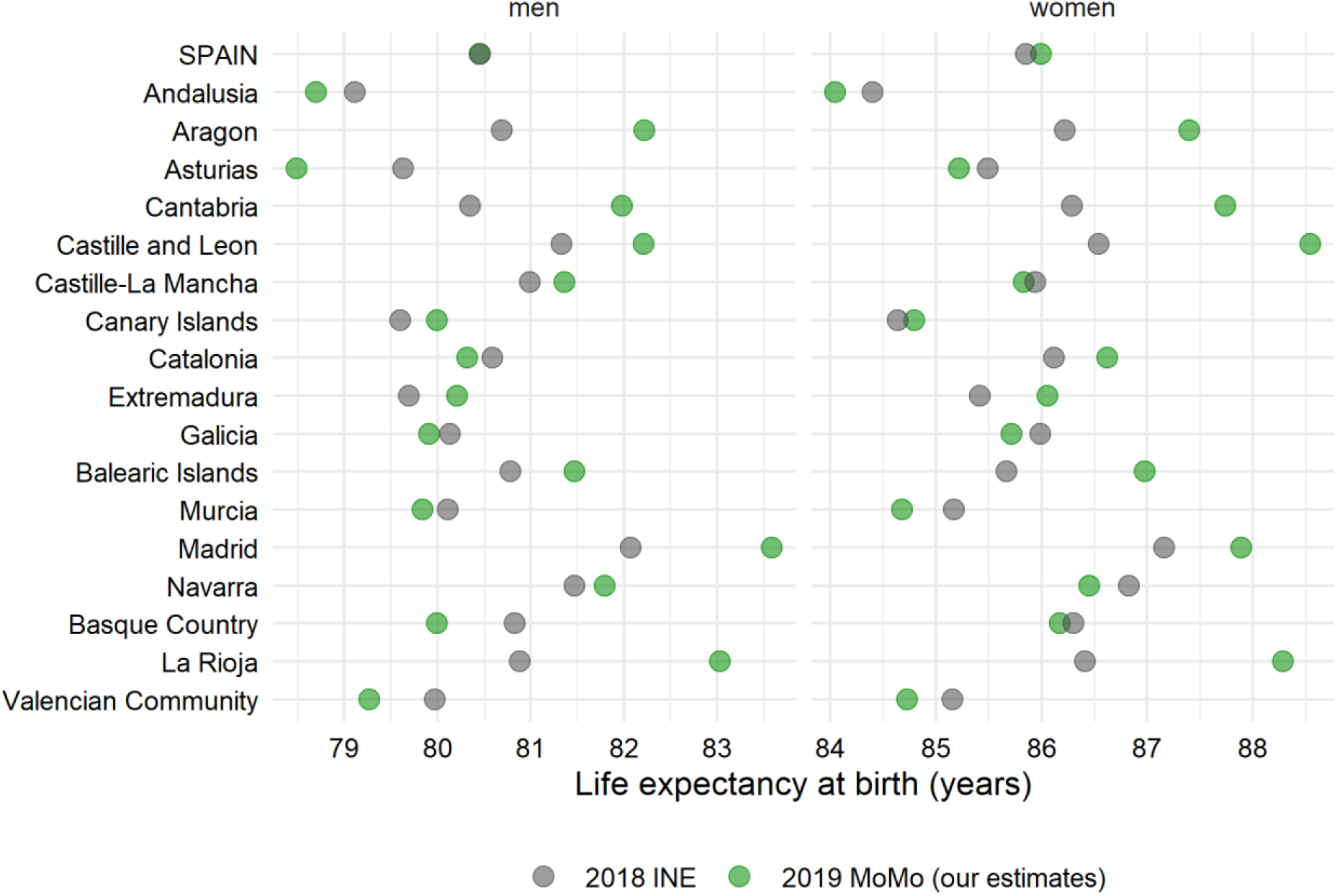
Annual life expectancy at birth in 2018 (INE) and in 2019 based on our estimates (MoMo data)

**Figure 3.**
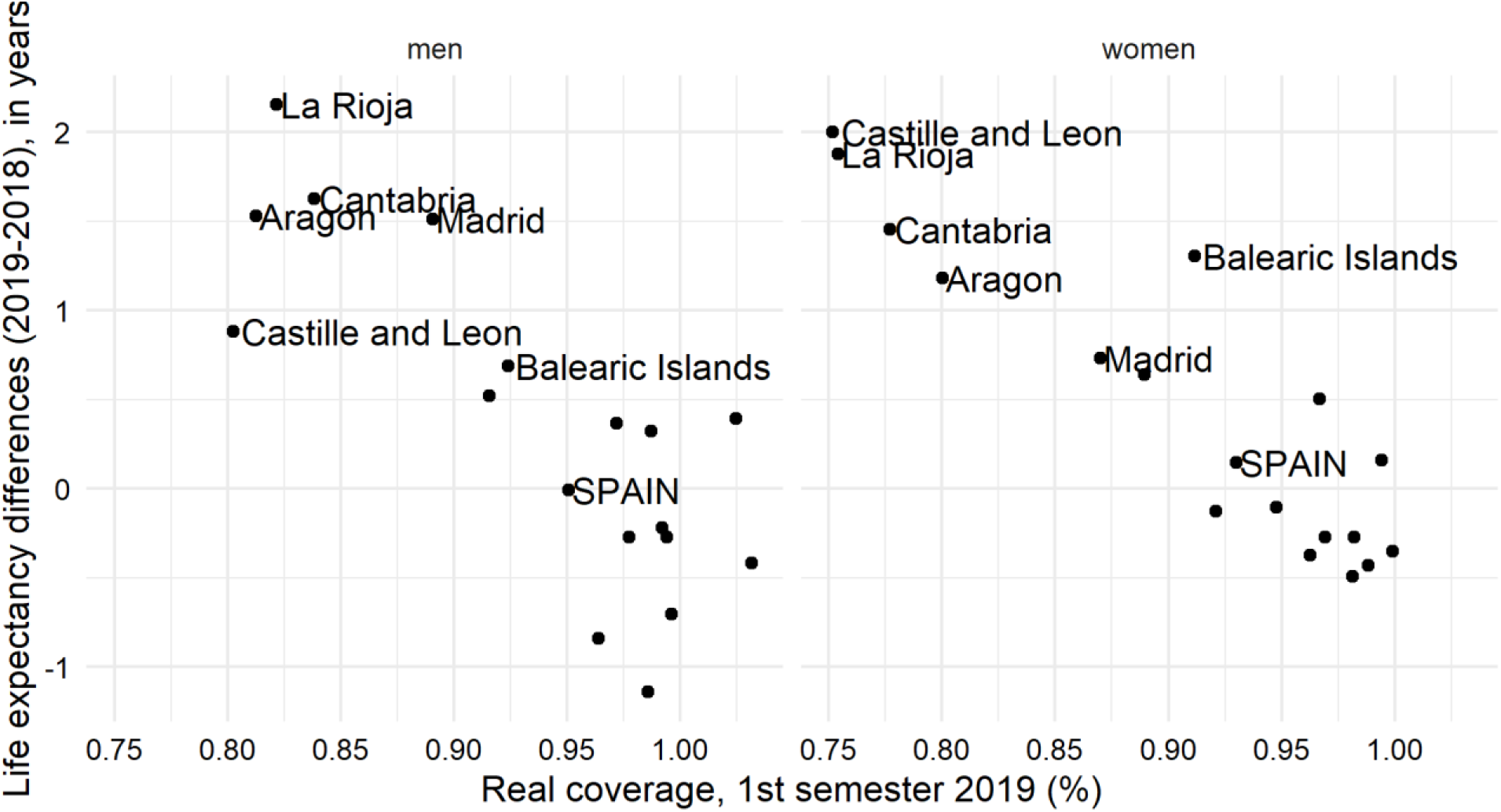
Associations between annual life expectancy at birth decline between 2018 (INE) and 2019 (own estimates, MoMo data) and coverage in MoMo

**Figure 4.**
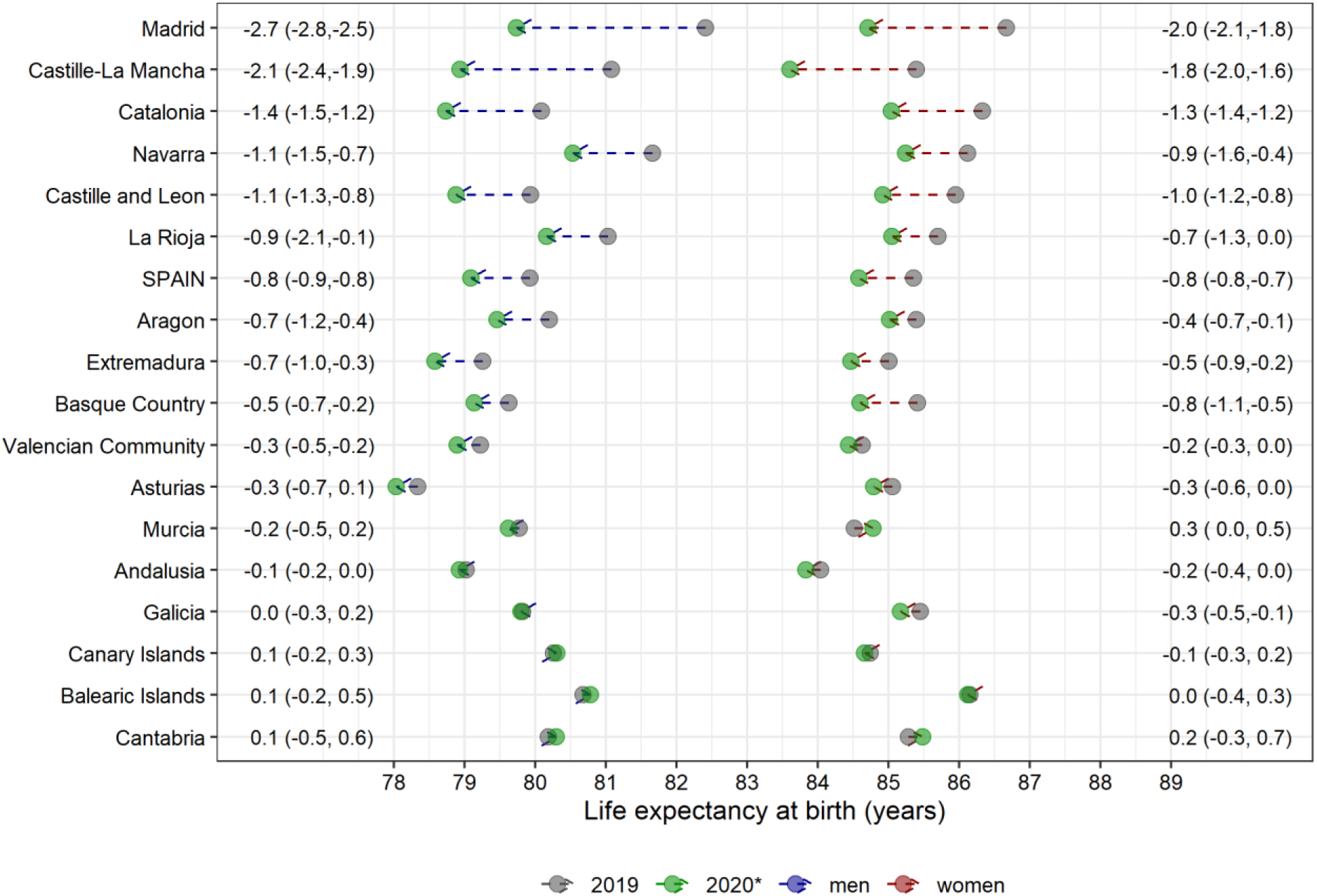
Annual life expectancy at birth in 2019, 2020* and differences between periods for Spain and its 17 regions by sex. Death counts corrected for the real coverage of MoMo* *1st semester of 2019. Correction made using provisional death counts from Movimiento Natural de la Población (INE)

**Figure 5.**
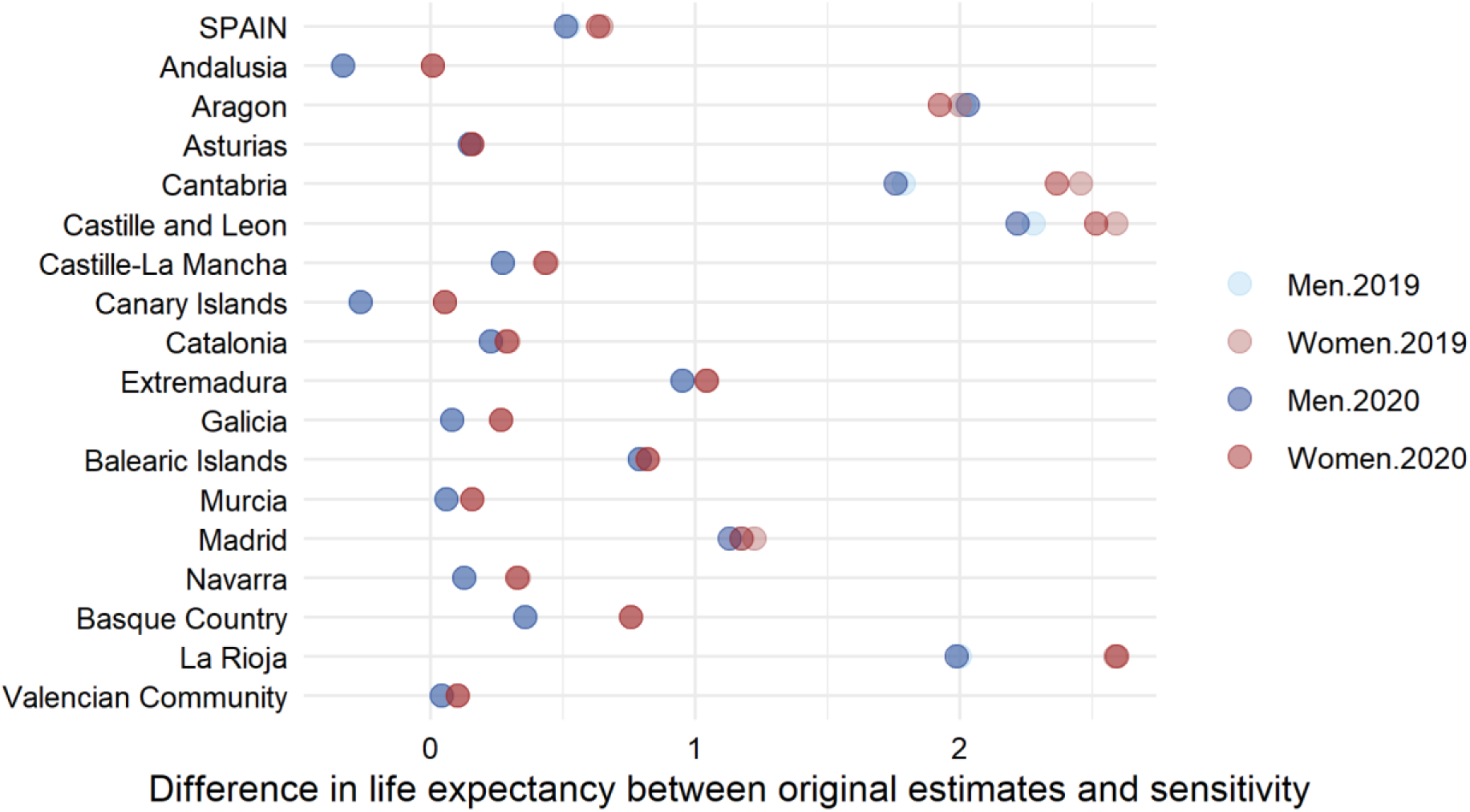
Differences in life expectancy at birth between our original estimates and the sensitivity analyses where death counts are corrected for the real coverage of MoMo

